# The quality of condition suggestions and urgency advice provided by the Ada symptom assessment app assessed with independently generated vignettes optimized for Australia

**DOI:** 10.1101/2020.06.16.20132845

**Authors:** Stephen Gilbert, Shubhanan Upadhyay, Paul Wicks, Claire Novorol

## Abstract

**Objectives:** To investigate the quality of condition suggestion and urgency advice provided by the Ada symptom assessment application and to compare it to published literature evaluation of free website and mobile symptom assessment applications accessible in Australia.

**Design:** The application was tested with 48 independently created medical condition vignettes using an externally developed methodology.

**Main outcome measures:** Correct condition suggestion (provided in first, the top three or top ten condition suggestion results); correct level of urgency advice (appropriate urgency category recommended).

**Results:** The correct condition suggestion was listed first in 65% of vignettes, and included among the first three results in 83% of vignettes. Urgency advice was exactly matching the vignette gold standard in 63% of cases, including about 67% of emergency and urgent cases and 57% of less serious case vignettes.

**Conclusions:** This study provides an analysis of the performance of one of the most used symptom assessment applications in Australia, the Ada app, which had not previously been evaluated in the literature in an Australia specific context. The app’s accuracy of condition suggestion and its provision of appropriate urgency advice is higher than of other symptom assessment apps evaluated in this context in the literature. We strive for continual improvement to ensure the most appropriate local advice on conditions and care.

Article Summary

Strengths and limitations of this study
This study addresses the performance of the Ada symptom assessment application in terms of condition suggestion accuracy and urgency advice accuracy, using fully independently created vignettes optimised for the Australian setting. Although the vignettes primarily described simple scenarios in which the patients did not have comorbidities, this study none-the-less provides a clear and structured assessment of the Ada app and allows direct comparison between the performance of the Ada app and that of other apps commonly used in Australia.

## Introduction

As summarised in [1] and shown in [2], Australians are generally embracing of online and smartphone technologies, and a high proportion of adults have access to the internet and own smartphones. The enthusiasm of Australians for these technologies also extends to their searching for information about healthcare, and about 80% of Australians report searching the internet for health information [3]. Symptom assessment applications are algorithm-based Smartphone and internet programs that ask patients questions about their demographic, relevant medical history, symptoms, and presentation. In the first few screening questions some symptom assessment apps exclude patients from proceeding if they are too young, too old, are pregnant, or have certain comorbidities. Assuming the user is not excluded, these software tools use a range of algorithmic approaches to suggest one or more conditions that might explain the symptoms (e.g. common cold vs pneumonia). Many symptom assessment apps then suggest next steps that patients should take (levels of urgency advice, e.g. self-care at home vs seek urgent consultation), often along with evidence-based condition information for the user.

Semigran and colleagues tested 23 free online symptom assessment applications with standardised patient vignettes in 2015 and compared their performance to general practitioners in 2016 [4]. They found that the symptom assessment applications provided the correct condition suggestion in 34% of evaluations, and appropriate urgency advice was provided in 57% of cases [5]. These were important studies, but they are now relatively outdated compared to the rapidly advancing field of symptom assessment apps, and many of the most widely used apps today were not investigated by Semigran et al.

The study of Hill et al. published in 2020 [1] is an update to the approach of Semigran et al. [5], and it was designed to evaluate the most prominent freely available apps in Australia. The vignettes included scenarios that health providers commonly encounter in Australia, including shingles, heart attacks, and viral upper respiratory infections [6].

In the Hill et al. study, apps were selected using criteria to identify those most prominent in internet search engines and app stores, with the process guided by structured advice in [7]. However, as recognised in [7], “ app stores are challenging to navigate, so it is important to fine-tune and filter app searches with the most relevant and targeted keywords”. App store searching in [1] was carried out using the terms “ symptom checker” and “ medical diagnosis”, “ health symptom diagnosis” and “ symptom”. However, the use of search terms including “ diagnosis” can exclude many symptom assessment apps, as those that are available in Europe are regulated and must be CE-marked, and will not describe themselves as diagnostic tools, as diagnosis is a function carried out by a doctor. The CE-marked Ada health assessment app was not identified or selected by [1], despite being freely available in Australia since 2016 [8], and despite being downloaded at least 200 times more in Australia in the period of November 2018 - January 2019 than either Symptomate or Symcat (data source appannie.com download data, accessed 13 June 2020) [9].

Therefore, the aim of our current study was to extend the findings of Hill et al. by investigating the condition suggestion and urgency advice performance of the Ada app with the same set of vignettes. This would allow greater context for the results from 36 other free-to-Australian-user symptom assessment applications, as already evaluated by Hill et al [1].

## Methods

### Symptom Assessment application

One software/application was assessed in this study: the Ada symptom assessment application. The justification for the selection of this application is provided in detail in the introduction - briefly, the Ada app is long established and highly used in Australia, and therefore it is a useful addition to the literature to assess the app with the 48 independent vignettes developed by [1]. The Ada app is evaluated in this paper using the same methods as reported in [1]. It could be that some other symptom assessment applications, popular in Australia, were not identified in the search protocol in [1] - this study has a narrow scope, which is the assessment of the Ada application only with the independent vignettes.

### Patient vignettes

The vignettes used in this study are exactly those created by [1]. Briefly, 30 patient vignettes from the study by Semigran and colleagues [4] were selected and adapted by [1] and were supplemented with 18 new symptom-based scenarios, designed with reference to the scientific literature, which were designed to include several reflecting Australia-specific conditions. The urgency advice of the vignettes was allocated into four triage categories: emergency, urgent, non-urgent, and self-care. The Ada app’s 8 urgency advice levels can be mapped to these advice levels, as shown in **Table 1**. This mapping is identical to the mapping used in [1] for the evaluated symptom assessment applications.

**Table 1.** Mapping of the Ada app’s levels of urgency advice to the gold standard triage categories from [1].

| Ada level # | Ada advice level | Gold standard vignette triage advice |
| --- | --- | --- |
| 1 | call ambulance | Requires emergency care |
| 2 | go to emergency department |  |
| 3 | see primary care within 4hrs | Requires urgent care |
| 4 | see primary care same day |  |
| 5 | see primary care 2-3 days | Non-urgent care reasonable |
| 6 | see primary care 2-3 weeks |  |
| 7 | do self-care / see pharmacist | Self-care reasonable |
| 8 | do self-care |  |

The medical conditions covered by the 48 vignettes can be classified as “ common” (85% of vignettes) or “ uncommon” (15% of the vignettes). This provides a good representation of general practice presentations in Australia, where 10% of presentations relate to uncommon conditions [10]. The full vignettes are listed in the data supplement of [1] and the vignettes are summarised in **Supplemental Table 1**.

The lay language summaries of the vignettes and the identified primary complaint were used for the entry of the vignettes in the Ada app. To maintain consistency, one investigator entered the information for each vignette (coauthor S.G). S.G. has not been involved in the development of Ada’s medical intelligence, question flow or interface design. The vignettes were entered between 12 June 2020 and 14 June 2020 using an Android Smartphone and using version 3.5.0 of the Ada symptom assessment application as it is available on the Australian Android (Google) app store. The Ada app has a broad coverage of user populations (e.g. childhood conditions, conditions in pregnant women), and it was therefore possible to enter all 48 vignettes.

### Symptom checker performance: condition suggestion

The Ada app provides the user with between one and five condition suggestions at the end of each symptom assessment. ‘Accurate condition-suggestion’ was defined as including the gold standard diagnosis as the top result (top-1), or as being among the top three (top-3) or top ten (top-10). In this paper, the top-10 potential condition-suggestions are listed, to allow for easy comparison to the websites and apps evaluated in [1], but as the Ada app provides a maximum of five condition suggestions top-10 is always equal to top-5. “ Incorrect condition suggestion” was defined as the correct condition not being included in the top-5 results. The decision of whether or not the condition suggested by the Ada app was a match for the gold standard diagnosis was made by coauthor S.U., who has over 5 years of primary care and emergency department clinical experience. Strict matching criteria were applied - the condition provided by the Ada app must have fallen into the set listed for the gold standard diagnosis by [1], with alternative medical names for the same condition being allowed.

#### Symptom checker performance: urgency advice

Urgency advice accuracy was defined as provision of an level of urgency advice which matched the gold standard vignetted triage, as defined by [1]. The Ada app always provides a single overall urgency advice for the symptom assessment, so an unambiguous evaluation was possible for each vignette.

### Data analysis

Simple descriptive statistical methods have been used to report the performance of the Ada app in the same format as the other apps assessed in [1].

#### Patient and public involvement

Patients were not involved in setting the research questions, the design, outcome measures or implementation of the study. They were not asked to advise on interpretation or writing up of results. No patients were advised on dissemination of the study or its main results.

## Results

### Condition suggestion performance

The correct condition suggestion was listed first in 65% of vignettes, and included among the first three results in 83% of vignettes. The condition suggestion results are summarised in **Table 2** and the complete solution provided by the Ada app for the full set of vignettes is shown in **Supplemental Table 1**.

**Table 2.**
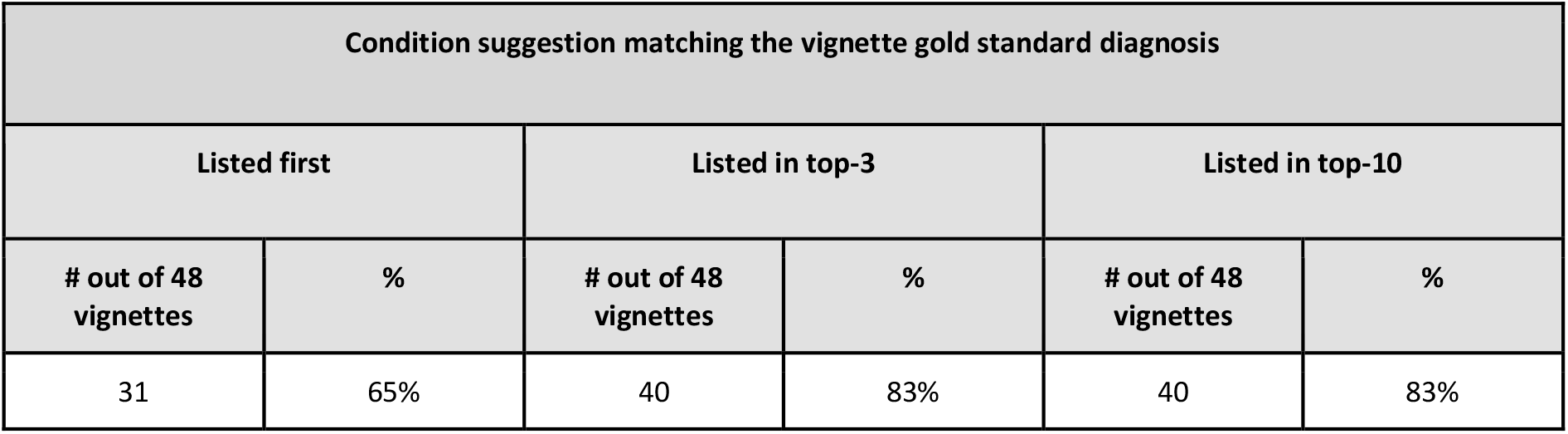
Accuracy of the condition suggestions provided by the Ada app. The data in this table can be compared with Supplementary Table 4 in [1] to provide context to the symptom assessment applications evaluated in that study. Note: (1) the Ada app provides a maximum of five condition suggestions, but the ‘Listed in top 10’ column is provided for ease of comparison to [1]; (2) In this study, the Ada app’s performance, as shown in the ‘Listed in top 10’ column is identical to the performance in the ‘Listed in top 3’ column, as all gold standard diagnosis identified were in Ada’s top-3 condition suggestions.

| Condition suggestion matching the vignette gold standard diagnosis |  |  |  |  |  |
| --- | --- | --- | --- | --- | --- |
| Listed first |  | Listed in top-3 |  | Listed in top-10 |  |
| # out of 48 vignettes | % | # out of 48 vignettes | % | # out of 48 vignettes | % |
| 31 | 65% | 40 | 83% | 40 | 83% |

### Urgency advice performance

Urgency advice was exactly matching the vignette gold standard in 63% of cases, including about 67% of emergency and urgent cases and 57% of less serious case vignettes. The urgency advice results are summarised in **Table 3** and the complete solution provided by the Ada app for the full set of vignettes is shown in **Supplemental Table 1**.

**Table 3.** Accuracy of the urgency advice levels provided by the Ada app. The data in this table can be compared with Supplementary Table 6 in [1] to provide context to the symptom assessment applications evaluated in that study.

| Levels of Ada app urgency advice matching the vignette gold standard triage |  |  |  |  |  |  |  |  |  |
| --- | --- | --- | --- | --- | --- | --- | --- | --- | --- |
| All cases |  | Emergency care required |  | Urgent care required |  | Non-urgent care reasonable |  | Self-care reasonable |  |
| # out of 48 vignettes | % | # out of 13 vignettes | % | # out of 14 vignettes | % | # out of 11 vignettes | % | # out of 10 vignettes | % |
| 30 | 63% | 12 | 92% | 6 | 43% | 6 | 55% | 6 | 60% |

## Discussion

### Principal findings

It was possible to enter 100% of the vignettes into the Ada app. The Ada app provided the correct condition as a first suggestion (top-1) in 65% of vignettes (compared to 10%-52% in [1]) and as a suggestion in the top-3 in 83% of vignettes (compared to 21%-69% in [1]). The best performing app reviewed by Hill et al. performed 13% less well than the Ada app in top-1 and 14% less well in the top-3 condition suggestions. In Hill et al’s original study [1], data was presented for a ‘provided answer’ approach, where the condition suggestion accuracy of symptom assessment apps were evaluated on the basis of the number of vignettes for which a condition suggestion was provided. In this analysis approach, the Ada app’s performance is identical to the ‘required answer’ analysis as all vignettes could be answered. However using the ‘provided answer’ approach the correct first condition suggestions (top-1) in [1] ranged between 12%-61% and for top-3 correct condition suggestions between 23%-77%. With this more generous scoring system the best performing app performed 4% less well than the Ada app in top-1 and 6% less well in the top-3 condition suggestions.

The urgency advice of the Ada app exactly matched the vignette gold standard in 63% of all cases. The range of performance of the apps evaluated in [1] which provided any urgency advice was 13%-58%, when considered from a ‘required-answer’ standpoint or 17%-61% with a ‘provided-answer’ approach. The urgency advice of the Ada app exactly matched the vignette gold standard in 67% of emergency and urgent cases and 57% of less serious case vignettes. This compared to advice that was matching in 49% of all cases, including 60% of emergency and urgent cases but only 30–40% of less serious case vignettes for the apps evaluated in [1]. A detailed breakdown of relative performance of each app evaluated in [1] is not provided here as this can be seen in Supplementary Table 6 of that publication.

In comparing the urgency advice performance of each app from that study, it should be taken into consideration that the urgency advice provided by symptom assessment apps needs to be evaluated across the whole spectrum from ‘emergency care required’ to ‘self-care reasonable’ for each app, as it is relatively easy to score well in one category at the expense of performance in the other categories.

### Comparisons to the wider literature

By design, the results of the current study can be compared to Hill et al [1] in detail, and this has been done in the section above. However it is also interesting to look at the results in the context of the 2016 study of Semigran et al. [5] upon which Hill et al’s [1] was based. The range of app condition suggestion performance in [5] was 4% to 44% for first condition suggestion (for the required-answer approach). In [1], four apps performed better than the best performing app in [5], and the best performing app performed 6% better. The Ada app performed 21% better than the best performing app in [4]. The range of app condition suggestion performance in [5] was 16% to 71% for top three condition suggestions (for the required-answer approach). With respect to the top-3 conditions suggested, in [1], four apps performed better and the best performing app performed 1% better. The Ada app performed 12% better than the best performing app in [4]. There does not appear to be a remarkable improvement between top-3 performance in [5], and top-3 performance in [1], but there are a number of explanations for this. One explanation is that most of the best performing apps in [1] have been developed for worldwide use, and may not have been particularly tuned in their medical content or advice for the Australian setting. A second explanation could be that the vignettes added to [1] are for relatively rare conditions, are relatively harder in their presentation, or are more tricky in their presentation that the original vignettes in [5]. The inclusion of the uncommon conditions Hendra virus, Queensland tick typhus and Ross River virus in [1] (see the full set of vignettes in **Supplemental Table 1**) suggests these vignettes may indeed be harder. Also, the inclusion of blue bottle Jellyfish sting and nettle sting, which are conditions where the user is highly likely to already know the cause before searching, could be seen as trickier for symptom assessment apps. The inclusion of these vignettes in [1] is entirely valid and very relevant to the Australian context, but it has the consequence that direct comparison of app performance between [1] and [5] is challenging.

The finding of this study, that the Ada app has relatively high condition suggestion performance and urgency advice performance compared to other available symptom assessment applications, reflects the finding of other studies [11–14]. The Ada app was recently compared to general practitioners (GPs) and competitor apps in a 200 vignettes study [11]. Many symptom assessment evaluations have focused on a single symptom assessment app or specific medical subdiscipline/specialism with a small number of vignettes, so we carried out a new study, in collaboration with medical experts from Brown University and with advice from other academic medical experts including from the UCL Institute of Health Informatics, to look at a broader spectrum of applications. The study compared the condition coverage, accuracy, and safety of 8 popular symptom assessment apps (including the Ada app) with each other and with 7 General Practitioners. The results showed that the Ada app’s performance was closest to that of the human doctors; it offered 99% condition coverage for the vignettes, gave safe advice 97% of the time (the same performance as GPs), and provided the correct suggested conditions in its top 3 about 70% of the time.

### Implications for clinicians and policymakers

As described above, and partly as a consequence of being one of the most widely used apps, and its optimization based on user feedback, the Ada app performs well compared to the other apps assessed in [1]. It is important that the Australian medical community considers the findings of the current study alongside the findings of [1] and the other literature cited above, as the Ada app is one of the most widely used symptom assessment apps in Australia.

### Unanswered questions and future research

This study uses a relatively small number of vignettes which are relatively simple in structure. In future studies we will carry out country-specific vignettes studies which will address in more detail the coverage of local conditions, the accuracy and appropriateness of advice for the local setting, and a larger numbers of vignettes, including those with more complex comorbidity and disease progression detail. Although vignettes studies are a high-throughput method, they are not a substitute for studies involving real users. Studies are currently underway in Europe and the US with real users for this reason, with further studies planned in Africa and Eastern Europe. It is also a goal to conduct real patient studies relevant to Australia in the future.

### Strengths and limitations of this study

The vignettes primarily described simple scenarios in which the patients did not have comorbidities and few of the vignettes reflected the complexity of real patients. The investigator who entered the vignettes was familiar with the Ada symptom assessment app as a user/evaluator, however, he was not involved in the development of the app’s medical intelligence, question flow or user interface. The solution provided by the Ada app for the full set of vignettes can be referred to and verified in full, as this is provided in **Supplemental Table 1**. Notable strengths of the study were that it used fully independently created vignettes which were described in lay language by [1], and that the vignettes tested conditions specific to Australia. As this study was conducted using the same methodology and results as the used in [1], the performance of the Ada app can be compared to the 36 symptom assessment applications evaluated in that study. A strength of vignettes studies, including this study, is that systematic comparisons between approaches or digital tools can be carried out.

## Conclusions

This study provides an analysis of the performance of one of the most used symptom assessment applications in Australia, the Ada app, which had not previously been evaluated in the literature in an Australia specific context. The app’s accuracy of condition suggestion and its provision of appropriate urgency advice is higher than of other symptom assessment apps evaluated in this context in the literature. We strive for continual improvement to ensure the most appropriate local advice on conditions and care.

## Data Availability

All data relevant to the study are included in the article or uploaded as supplementary information.

## Footnotes

### Author Statement

S.G. and C.N. contributed to the planning (study conception, protocol development). S.G. entered the vignettes in the Ada app. SU adjudicated if the conditions suggestions matched the vignette gold standard diagnoses, S.G. carried out the data analysis & interpretation. P.W. contributed to the reporting (report writing). All the authors contributed to commenting on drafts of the report. S.G. is the guarantor for this work. The corresponding author attests that all listed authors meet authorship criteria and that no others meeting the criteria have been omitted.

### Disclaimer

None

### Patient consent for publication

Not relevant - no direct patient data.

### Provenance and peer review

Not commissioned; externally peer reviewed.

### Supporting Information

Additional Supporting Information is included with the online version of this article.

